# Discriminative Subtyping of Lung Cancers from Histopathology Images via Contextual Deep Learning

**DOI:** 10.1101/2020.06.25.20140053

**Authors:** Benjamin J. Lengerich, Maruan Al-Shedivat, Amir Alavi, Jennifer Williams, Sami Labbaki, Eric P. Xing

**Affiliations:** Carnegie Mellon University; Mohamed bin Zayed University of Artificial Intelligence

## Abstract

Summarizing multiple data modalities into a parsimonious cancer “subtype” is difficult because the most informative representation of each patient’s disease is not observed. We propose to model these latent summaries as *discriminative subtypes*: sample representations which induce accurate and interpretable sample-specific models for downstream predictions. In this way, discriminative subtypes, which are shared between data modalities, can be estimated from one data modality and optimized according to the predictions induced in another modality. We apply this approach to lung cancer by training a deep neural network to predict discriminative subtypes from histopathology images, and use these predicted subtypes to generate models which classify adenocarcinoma, squamous cell carcinoma, and healthy tissue based on transcriptomic signatures. In this way, we optimize the latent discriminative subtypes through induced prediction loss, and the discriminative subtypes are interpreted with standard interpretation of transcriptomic predictive models. Our framework achieves state-of-the-art classification accuracy (F1-score of 0.97) and identifies discriminative subtypes which link histopathology images to transcriptomic explanations without requiring pre-specification of morphological patterns or transcriptomic processes.

## Introduction

Cancer is a heterogeneous disease which can be phenotypically variable even for tumors of a single tissue [PSE^+^00, SPT^+^01, FPS13]. For this reason, subtype discovery — the classification of tumors into biologically- and clinically-relevant groups — is a crucial task to advance the understanding and treatment of cancers. A common approach is to define *molecular subtypes* with a small number of marker genes [vdVHvV^+^02, RVG^+^11, CCL^+^08, CBCB19, BKH^+^02] identified through immunohistochemistry or gene expression analysis. Characterization of these molecular subtypes is continuously advancing, and much recent work has focused on identifying fine-grained descriptions which comprise previously-characterized subgroups [MBW^+^13, KMK^+^13, RPB^+^19, CGAR12, WYH^+^10]. While subtypes are continuously being refined through experimentation, a fundamental question remains: how would we know when the optimal cancer subtypes have been defined?

This question inspires us to propose a machine-learning based criterion for optimality of subtyping classifications: a set of subtypes is optimal if it most concisely describes cancers in such a way that downstream predictions require minimal extra information to be maximally accurate. We call these optimal subtypes *discriminative subtypes* and show that they can be estimated through contextual machine learning. In lung cancers, these discriminative subtypes correspond to distinct transcriptomic and morphological patterns, suggesting that different tumors should be understood in light of different pathways.

### Contributions

We examine three hypotheses surrounding this task of distinguishing LUAD and LUSC tumors: (1) there exist “discriminative subtypes” of lung cancers which improve downstream predictive models, (2) these subtypes can be predicted from histopathology images via contextual deep learning, and (3) the discriminative subtypes correspond to biological processes and morphological patterns. The validation of these hypotheses suggests that macroscopic morphological patterns can be used to predict intra-cellular genomic profiles which predict outcomes, even when the genomic profiles are unknown *a priori*. Beyond lung cancer, discriminative subtypes and contextual deep learning can improve biomedical understanding of cancer subtypes by connecting histopathology images to gene expression patterns in a new end-to-end, fully-differentiable manner which does not require pre-specifying which transcriptomic patterns are of interest.

### Discriminative Subtypes

We propose a *discriminative subtype* (Figure 1) as a latent variable which captures the variation in many observable variables, and optimally “contextualizes” predictive tasks by providing a compressed view of relevant information. Discriminative subtypes may correspond to, but are not limited to, previously-characterized molecular subtypes. The optimal descriptions of tumors are multi-faceted; consequently, we would like to automatically identify contexts that summarize the key information of each sample rather than pre-define molecular subtypes.

**Figure 1:**
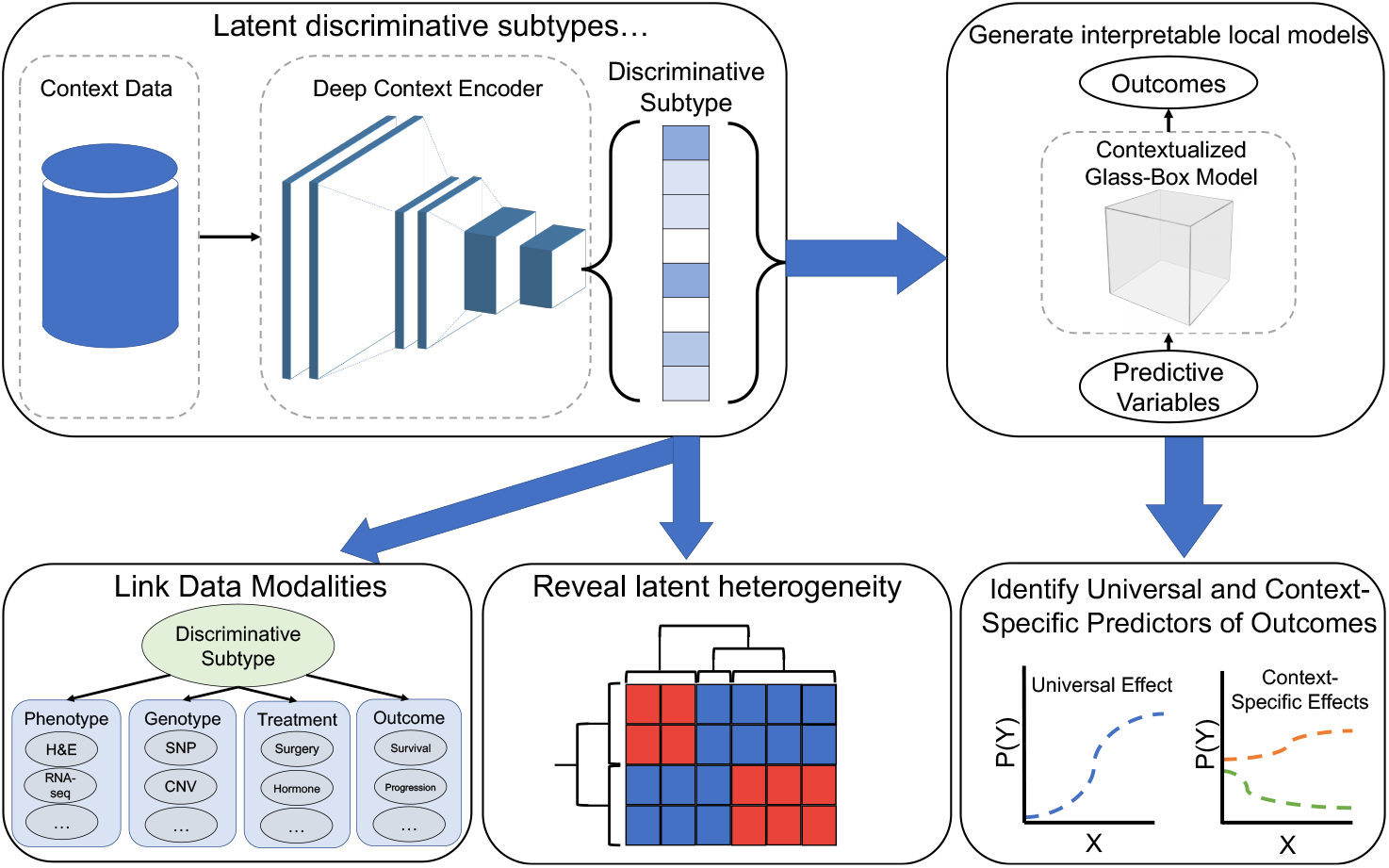
We propose that the most concise description of the many variables regarding oncology patients is an unobserved latent variable, and we seek to estimate this latent *discriminative subtype* which improves the discriminative ability of downstream glass-box predictors. With the optimality of these discriminative subtypes defined by the predictive ability of downstream models, they can be optimized through end-to-end contextual machine learning without pre-specifying any subtypes or factors. This links data modalities, reveals latent heterogeneity, and identifies universal and context-specific predictors of outcomes.

#### Discriminative Subtyping of Lung Cancers

Lung cancer is common, accounting for 13% of all new cancer cases and over 20% of all cancer deaths [SMJ20]. Lung adenocarcinoma (LUAD) and Lung Squamous Cell Carcinoma (LUSC) are two of the most common subtypes [CTM11] and have important distinctions in clinicopathology [RTGA19], immune subtypes [SKSK18], and therapeutic effectiveness [LCB^+^13], so distinguishing LUAD from LUSC is an important clinical task.

However, there may not be a single transcriptomic signature which universally distinguishes LUAD/LUSC. Instead, we would like to allow different cellular processes to predict subtype for different samples. To do this, we use a deep neural network to summarize histopathology “contexts” into discriminative subtype of each sample, and then use this latent discriminative subtype to generate sample-specific coefficients for a logistic regression model which predicts LUAD/LUSC from gene expression. This approach summarizes biological processes as interpretable coefficients of genes, and connects these biological processes to histopathology morphology through latent discriminative subtypes.

As a result, discriminative subtypes go beyond previous studies which have used morphology to estimate the likelihood of individual mutations [ADME18] or gene expression [FJT^+^20, ZFZM20] (more complete related work is described in Section A); instead, discriminative subtypes are a data-driven approach to learn gene expression patterns that are meaningful in morphological contexts and to summarize biological patterns on a process level. We investigate the utility of this approach to predict subtypes from the histopathology images and transcriptomic data of patients diagnosed with LUAD or LUSC. This approach not only achieves a new state-of-the-art accuracy for LUAD/LUSC classification, but also provides parsimonious descriptions that link morphology and transcriptomics.

## Results

Our methodology (detailed in Materials and Methods) uses histopathology images to contextualize transcriptomic models by predicting the discriminative subtype of each sample. This procedure not only improves the accuracy of predictions made by the transcriptomic models, but also provides transcriptomic “archetypes” which can be summarized as biological processes and traced to morphological patterns. This goes beyond previous studies which have used morphology to estimate the likelihood of individual mutations [ADME18] or gene expression [FJT^+^20, ZFZM20]; here, we allow the model to learn which gene expression patterns are meaningful in each morphological context, allowing us to summarize the biological patterns and tie them to morphology on a process level rather than a mutation level. The goal of this approach is not only to enhance the classification performance, but also to infer meaningful clusters. We first measure predictive accuracy and then we seek to assign biological meaning to the archetypes.

### Contextualization Improves Prediction of Subtypes

First, we measure the ability of the model to classify LUAD, LUSC, and healthy samples. Our model (CEN) achieves the best classification results, outperforming all the multi-modal and the uni-modal baselines to achieve a new state-of-the-art performance for classifying LUAD/LUSC/Healthy samples (Tables 1, C.1). The InceptionV3 deep neural network was used by Coudray et al [COS^+^18] to achieve state-of-the-art classification of LUAD/LUSC samples from histopathology images, and our use of this model reproduces the performance reported therein. While this performance is state-of-the-art for histopathology data, a regularized logistic regression model operating on transcriptomic data significantly outperforms the deep neural network, indicating that the transcriptomic assay is a very good source of information regarding tumor subtype. By using both the transcriptomic and histopathology data, the multi-modal models outperform either single modality, and the CEN achieves the highest accuracy of all.

**Table 1:** Performance of models on 3-way Normal/LUAD/LUSC classification. We report accuracy on both the “patch” level (where each prediction corresponds to a small patch in the image) and on the “sample” level (where a single prediction is made for the whole-slide image). Some models are designed to operate on transcriptomic data (T), while some operate on histopathology data (H), while others use both forms of data (H+T). CEN (our model) outperforms all existing models.

| Level | Model | Data | Accuracy (%) | Macro F1 |
| --- | --- | --- | --- | --- |
| Patch | **CEN** | **H+T** | **96.18** | **96.97** |
|  | Concatenated | H+T | 95.32 | 93.65 |
|  | Ensemble | H+T | 94.61 | 90.23 |
|  | Logistic Regression | T | 94.05 | 91.40 |
|  | InceptionV3 | H | 69.14 | 65.85 |
| Sample | **CEN** | **H+T** | **94.51** | **95.29** |
|  | Concatenated | H+T | 93.33 | 93.65 |
|  | Ensemble | H+T | 92.94 | 90.23 |
|  | Logistic Regression | T | 92.16 | 89.67 |
|  | InceptionV3 | H | 80.00 | 76.76 |

### Archetypal Models Correspond to Distinct Biological Processes

The CEN model estimates 32 transcriptomic model archetypes which are used to predict LUAD/LUSC/Healthy labels in various discriminative subtypes. We seek to interpret these archetypes to understand the underlying biological processes. First, we ask whether the model archetypes are redundant or look at different biological processes. Surprisingly, the coefficient vectors of the archetypes are nearly orthogonal (Fig. C.10), indicating that archetypes emphasizes distinct processes.

Next, we use enrichment analysis [STM^+^05] to identify biological processes which are statistically over-represented in the archetype coefficients. For each archetype, we have 3 vectors of coefficients defining the 3-way (LUAD/LUSC/Healthy) logistic regression model. For each vector of coefficients, we sort the genes by the magnitude of the associated coefficient (top 5 genes for each model archetype for normal, LUAD, and LUSC labels are provided in the Supplement). We search the top 100 genes in this list for enriched biological terms against the background of the remaining 1595 genes using gProfiler [RAA^+^16]. We set a minimum intersection size of 3 genes (the number of genes selected from each term must be at least 3) and a maximum Bonferroni-corrected p-value of 0.05. Terms enriched in the archetypal models for prediction of the “Normal” label (which corresponds to case/control prediction) are shown in Table 2. Similar tables for the LUAD and LUSC models are available in the Supplement. Many of the 32 archetypal models are significantly enriched for markers of cancer, indicating that not only do the discriminative subtypes correspond to distinct *statistical* patterns but also distinct *oncologic* processes.

**Table 2:** Archetypal transcriptomic models with terms significantly enriched (*p <* 0.05).

| Arch. | Term ID | Term Name | P-Val |
| --- | --- | --- | --- |
| 1 | KEGG:04071 | Sphingolipid signaling pathway | 0.037 |
|  | KEGG:04310 | Wnt signaling pathway | 0.049 |
| 6 | REAC:R-HSA-6802952 | Signaling by BRAF and RAF fusions | 0.039 |
| 8 | TF:M06732 | Factor: ZNF304 | 0.023 |
| 12 | REAC:R-HSA-8939236 | RUNX1 | 0.022 |
| 13 | TF:M09657_1 | Factor: Smad4 | 0.004 |
| 15 | GO:0071385 | Cellular response to glucocorticoid stimulus | 0.013 |
| 17 | REAC:R-HSA-400206 | Regulation of lipid metabolism by PPARα | 0.018 |
| 18 | TF:M04726_1 | Factor: REST | 0.001 |
| 19 | TF:M05327_1 | Factor: WT1 | 0.025 |
| 21 | TF:M01224_1 | Factor: P50:RELA-P65 | 0.034 |
| 25 | TF:M09611_0 | Factor: ER81 | 0.003 |
| 26 | KEGG:04215 | Apoptosis | 0.006 |
| 28 | GO:0051240 | Pos. reg. of multicellular organismal process | 0.006 |
| 30 | REAC:R-HSA-4791275 | Signaling by WNT in cancer | 0.045 |

As a vignette, below we discuss the terms in Table 2 which have a p-value of less than 0.01:

- **13: Factor: Smad4**. SMAD family member 4 (SMAD4) is a necessary component of the transforming growth factor beta (TGF*β*) pathway and in this capacity regulates cellular proliferation [YDW^+^97, ZMD18, ZBZ^+^98]. SMAD4 has been established as a tumor suppressor in pancreatic, colon and lung cancer [HTK^+^16, MW12, ZMD18]. Decreased expression of SMAD4 has been identified as a marker of Non-Small Cell Lung Cancer and poor prognosis in LUAD [IBH^+^12, HTK^+^16, CSJ^+^17, MW12].
- **18: Factor: REST**. RE1-silencing transcription factor (REST) is most widely known to repress neuronal genes in non-neuronal cells, but has also been found to be an oncogene or tumor suppressor gene in certain cancers [NPDM13]. In breast cancer, it has been shown to act as a tumor suppressor, and loss of this gene is associated with aggressive breast cancer [WGS^+^10], while an isoform of REST has been indicated as a specific clinical marker for early detection of small cell lung cancers [CEWQ00, SSI^+^13].
- **26: Apoptosis**. A hallmark of cancer cells is the ability to limit or avoid cell death induced by apoptosis [HW11]. In lung cancer, multiple genes in the KEGG apoptosis pathway have been found to be correlated with tumorigenesis, chemoresistance, and poor prognosis [LPY^+^17, PHK13, yYKS^+^04, DLL^+^13, TTY^+^01, JEL^+^01, JEL^+^01, Fen05, GED^+^04]. For example, altered expression of caspases involved in the crucial final steps of apoptosis have been shown to make cells resistant to apoptosis, enabling resistance to chemotherapy [Fen05, PCS^+^12, JEL^+^01, TTY^+^01, yYKS^+^04].
- **28: Positive Regulation of Multicellular Organismal Process**. Oncogenes activate tumor growth and proliferation processes by up-regulating other development processes, including angiogenesis, cell migration, and metastasis. For example, aggressive cancers achieve metastasis by up-regulating inflammation and cell migration [HW00, HW11].

### Archetypal Models Correspond to Distinct Morphologic Patterns

One of the advantages of CEN is that it enables us to tie the morphologic patterns recognized in histopathology images into the biological processes of the transcriptomic models. Towards this end, we visualize representative patches which maximize the predicted influence of archetypal models 12, 13, and 30 in Figure 2 (similar results for the other archetypal models are shown in the Supplement). We can see that the morphology is more similar within the clusters corresponding to archetypes than between the clusters (e.g. archetypal model 12, corresponding to the RUNX1 factor, seems to focus on morphological features of hematopoietic processes and macrophage response, while archetypal model 13, corresponding to the SMAD4 intercellular signaling complex seems to focus on morphological features of irregular multicellular structure) indicating that the transcriptomic patterns used for accurate downstream predictions correspond with morphological changes.

**Figure 2:**
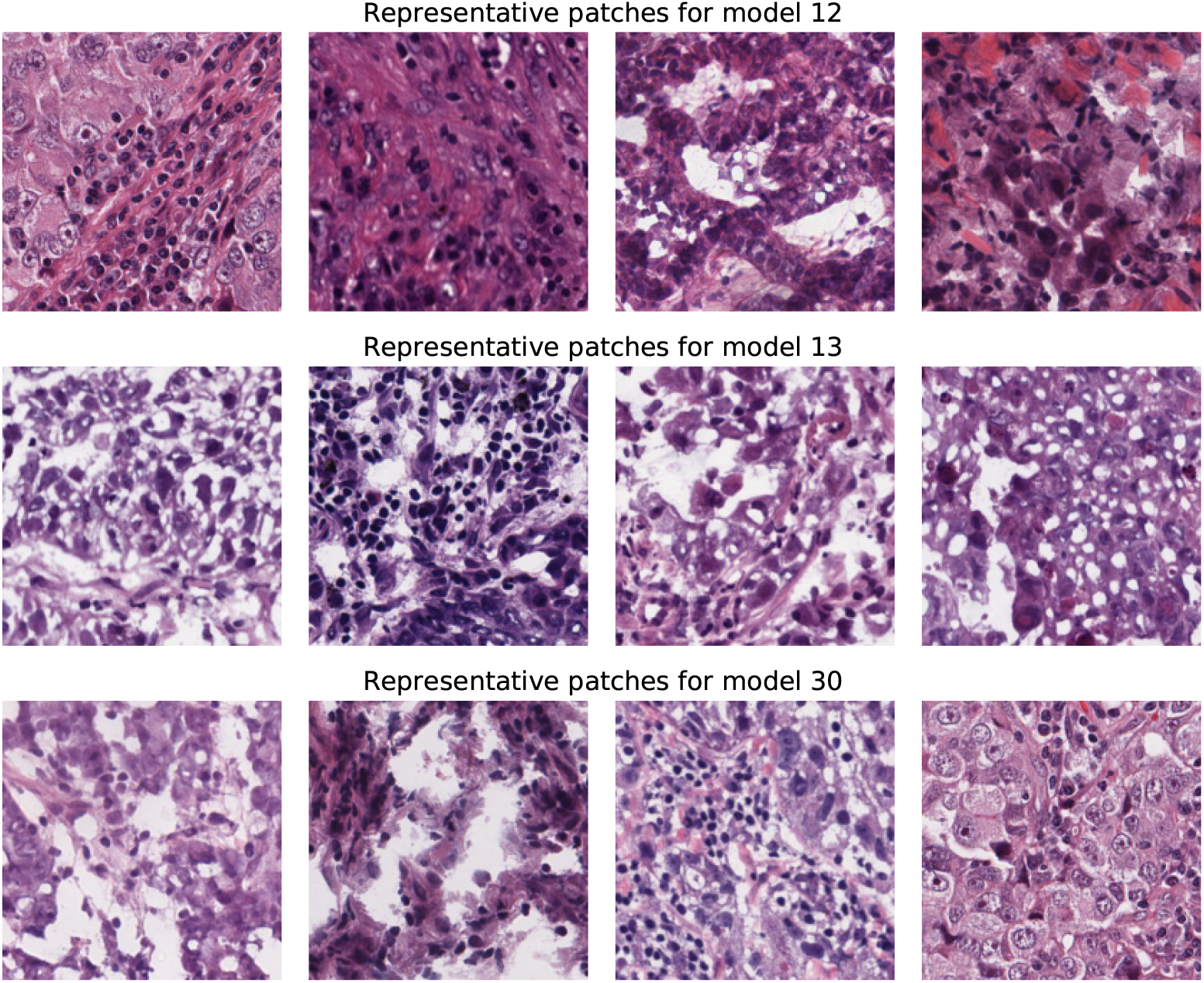
Morphology is homogeneous within each archetype and varies between archetypes. Each row contains the set of patches which made CEN assign the highest weights to the corresponding archetypal model over the entire training dataset. These patches represent archetypal morphological patterns for the archetypal transcriptomic models and link transcriptomic attention with morphology.

### Discriminative Subtypes Display Intratumor Heterogeneity

The spatial diversity of cells within lung tumors has recently been shown to encode prognostic markers [ARR^+^20]; the CEN model enables us to ask questions regarding the localization of discriminative subtypes. By making predictions for each patch independently, the CEN model allows us to ask whether different locations in the image are indicative of distinct tumor subtypes and if morphological patterns of a particular subtype tend to be clustered in a single location. Firstly, we see that on held-out test data, in 46.3% of patches, the CEN assigns highest weight to a discriminative subtype which is not the discriminative subtype chosen for the full slide by a majority vote of the patches. This indicates that there may either be significant diversity or uncertainty of morphological patterns or tumor subtypes in each tumor. To examine the spatial locality, we visualize the predictions of discriminative subtypes. Discriminative subtypes exhibit spatial locality (Figures 3 and C.1), with nearby patches tending to correspond to the same predicted subtype.

**Figure 3:**
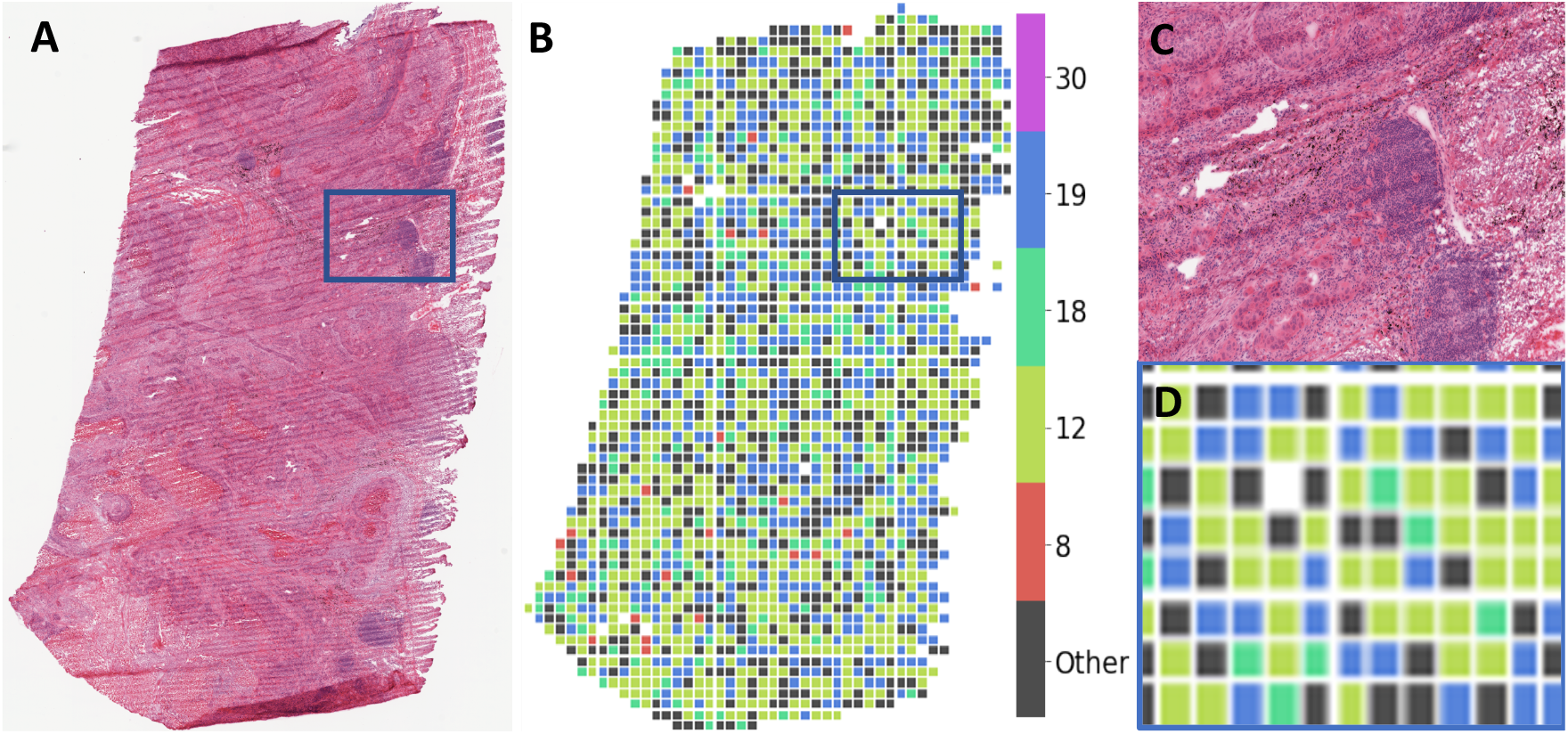
Discriminative subtypes display significant heterogeneity throughout the sample, but cluster spatially. Representative results of patch-level subtype prediction for a LUSC sample held out from CEN training. **(A)** tumor whole-slide image. **(B)** archetypes assigned to each patch by CEN; for visual clarity, we display only the common archetypes 8, 12, 18, 19, and 30, and group all other archetypes under “other”. **(C, D)** inset squares outlined in blue in panes A and B.

## Discussion

This work demonstrates the utility of estimating latent *discriminative subtypes* of lung cancers. We have defined discriminative subtypes as latent variables which provide parsimonious descriptions of a tumor, and these subtypes provide context which optimizes the ability of a downstream model to discriminate between subtypes of tumors. Contextualizing predictive models according to these discriminative subtypes achieves state-of-the-art classification accuracy for LUAD/LUSC/healthy prediction, suggesting that different transcriptomic patterns are most predictive in different contexts.

For machine learning practitioners, this has several implications. Firstly, multi-modal data analysis is critically important in healthcare (often because the technical noise in each measurement can most effectively be reduced by having multiple views of each sample), but multi-modal pipelines are difficult to meld with interpretability. Contextual learning is a promising framework for building interpretable multi-modal approaches. By allowing one data modality to select the important features for another modality, we can achieve both accuracy and interpretability.

This also has biomedical implications. Firstly, because the discriminative subtypes correspond to biological processes, we can surmise that these are distinctive signatures of cancers that may be used to improve personalized medicine beyond known histologic or molecular subtypes. Furthermore, because a deep learning model trained on hisotopathology images can assign meaningful contexts for transcriptomic models, some patterns of transcriptomic aberrations are contained in histopathology data with only basic hematoxylin and eosin (H&E) staining. This concords with several recent works to predict genetic variants from H&E stained histopathology data [PBC^+^16, ADME18, FJT^+^20], and encourages future works which discover and analyze morphological patterns of cellular modifications. All in all, this research demonstrates the utility of jointly modeling histopathology and transcriptomic data to identify latent cancer subtypes, and underscores the potential for histopathology images to be used for discovery of fine-grained subtypes.

## Materials and Methods

### Model Architecture

To combine multi-modal data in an interpretable pipeline, we use a version of Contextual Explanation Networks (CENs) [ASDX20] (Figure 4). The CEN architecture takes data represented by: (i) context variables *C*, (ii) interpretable variables *X*, and (iii) target variables *Y*. In this paper, we use histopathology images as contextual data and gene expression assays as interpretable predictors of the cancer type. The model represents conditional probability of the cancer type given the histopathology and transcriptomic inputs, ℙ(*Y* | *X, C*), in the following form:

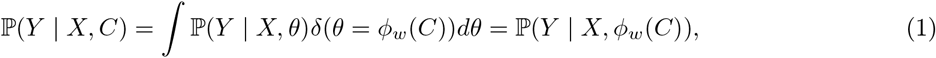

where ℙ(*Y* | *X, θ*) is a linear logistic model that predicts cancer types from gene expression. Note that parameters (or weights) *θ* of the logistic model are a function of the contextual information, i.e., *θ* = *ϕ*_*w*_(*C*). In other words, CEN produces a sample-specific parameterization of a linear probabilistic model that operates on transcriptomic data based on histopathology.

**Figure 4:**
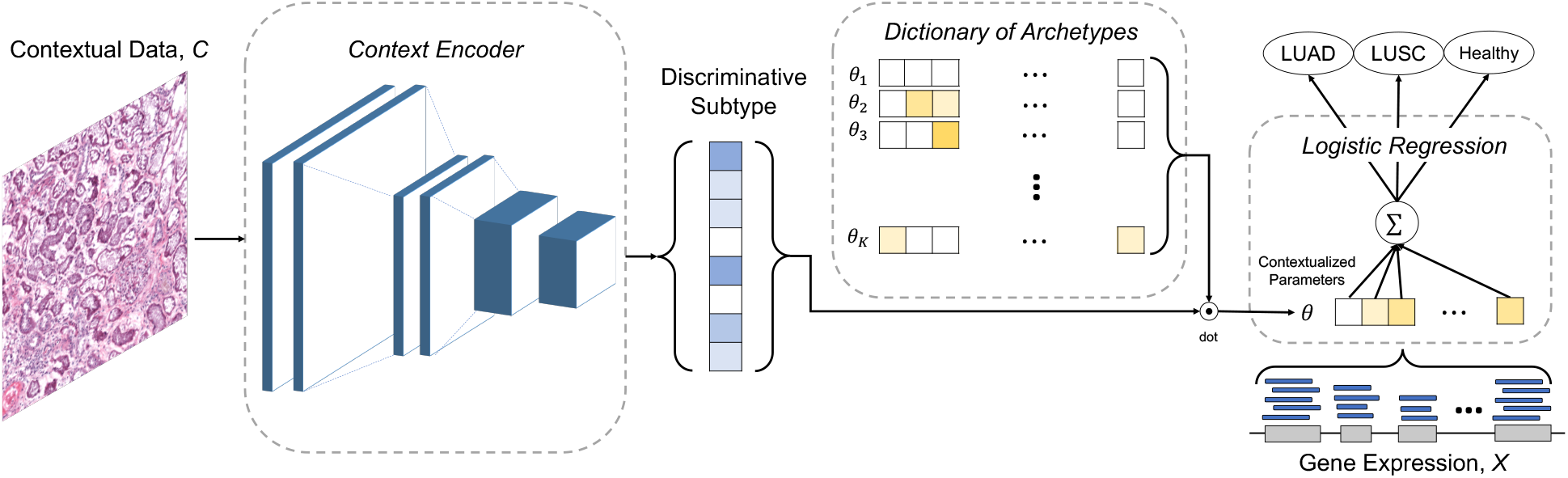
The CEN architecture estimates latent discriminative subtypes by combining “contextual” histopathology images (*C*) with gene expression (*X*) to predict sample classifications. The discriminative subtype is predicted from contextual data and forms sample-specific parameters *θ* by weighting archetypal models stored in a dictionary. The sample-specific logistic regression models estimate classifications from gene expression (*X*) with sample-specific coefficients, and this classification loss is back-propagated to update both the dictionary of archetypes and the context encoder.

Generation of parameters for sample-specific linear models is accomplished via a deep context encoder. Aligning with the hypothesis of discriminative subyptes, parameters *θ* are confined to be a linear combination of a small constant number (*K*) of “archetypes,” denoted {*θ*_1_, …, *θ*_*K*_ or *θ*_1:*K*_}. For a sample *i* with context *c*_*i*_, weights *θ*_*i*_ are:

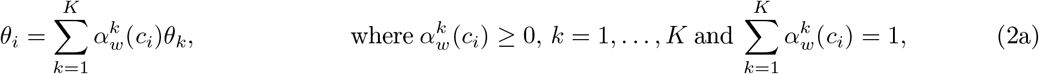

where (*α*^1^, …, *α*^*K*^) is a vector output of the context encoder, which we call the *discriminative subtype*. This dictionary of parameter sets, *θ*_1:*K*_, is estimated jointly with the context encoder, and the entire architecture is trained end-to-end via backpropagation after initialization of the archetypes to random vectors. To summarize, the context encoder is a deep neural network that processes contextual data (histopathology imagery) and outputs a probability vector of length *K* that softly selects weights for a linear probabilistic model from a dictionary of archetypes. Architectural decisions, hyperparameters, and baseline methods are available in Section B. Python code is available at: https://github.com/slabbaki/discriminative-subtyping.

### Cohort

We use lung cancer data available in the NCI Genomic Data Commons [GHF^+^16], which includes resources from both The Cancer Genome Atlas and The Cancer Imaging Atlas to provide multi-modal descriptions of a large number of cancer patients. As of April 2, 2020, this depository included 585 LUAD cases and 504 LUSC cases, along with a variety of cases with cancer of other tissues. From this set, we selected all samples which have both eosin-stained histopathology whole-slide images and transcriptomic RNA-seq profiling. This selects 992 patients, with 1 or 2 whole-slide images for each patient. We split this dataset by patient into training, validation, and testing partitions, so the test set consists of completely held-out patients (size and composition of partitions in Table B.1).

#### Image Subsampling

We use 20x magnification whole-slide histopathology images. To make these large images suitable for the pre-trained Inceptionv3 architecture, we split each whole-slide image into non-overlapping 299×299 image patches. The patches were controlled for quality by discarding any patches with more than 25% white pixels, following the procedure of [COS^+^18]. We treat each patch as a separate sample, with transcriptomic data duplicated over each patch.

#### Transcriptomic Profiles

The transcriptomic profiling in TCGA captures the expression of over 60,000 distinct transcripts in each sample. We select the 1000 transcripts with the highest variance in the non-lung cancer cases in TCGA. In addition, we augment this set with 695 transcripts corresponding to genes in the Catalogue of Somatic Mutations in Cancer (COSMIC, [BDF^+^04]).

## Data Availability

TCGA data is publicly available at the following link.

https://portal.gdc.cancer.gov/

## A Related Work

Molecular profiling has increased understanding of the pathology of cancer [RR17, TBB^+^15, BWSH^+^11, CBCB19], allowing pathologists to use phenotypic molecular and morphological data to identify increasingly specific cancer subtypes [Ina17, GWC^+^13, Sch10], but subtype identification has been hindered by the lack of knowledge of the biological processes underlying tumor growth [HW11, Daw17, HPL14, SSZF19]. In previous work, this was circumvented by using observed phenotypic and/or genotypic data to define cancer subtypes [KI18, SCM19, IKK^+^20, WTL^+^19, WCY^+^18, GSCM^+^19, CHG^+^19, MYA^+^18, MSC^+^19]. Similarly, rising interest in linking morphological patterns of cancer (e.g., histopathology images) with intra-cellular mechanisms [SGLGK22] has inspired many attempts to use deep learning to predict genetic mutations from morphology [FJT^+^20, VSS^+^17, ADME18, PBC^+^16]. While these approaches have proven that transcriptomic changes are predictable from detectable morphological patterns, these approaches are limited to pre-specified transcriptomic patterns. and the features identified capture only a subset of the biological processes underlying tumor growth and relate to only pre-specified tasks. Our work is the first to define cancer subtypes according to their implications on downstream discriminative models. This definition enables us to learn patient-specific models for downstream prediction tasks, linking data modalities and discovering meaningful patterns without pre-specification of genomic patterns.

### Challenges of Interpretable Multi-modal Predictions

A key challenge in multi-modal predictions is interpretability. Previous work [COS^+^18, WCY^+^18, GSCM^+^19, WTL^+^19, LZY^+^17, YZB^+^16, LdPM^+^19] built interpretable predictive models of only one modality, such as histologic patterns which are interpretable to pathologists. Multimodal models increase predictive capabilities, but interpretability has been hindered by the complexity of the models [ZYL^+^16, PBC^+^16, HKT^+^19, CZH^+^17, MYA^+^18]. The main way these models have been interpreted is through retrospective analyses of the features selected in their neural networks [HKT^+^19, MYA^+^18], and it is unclear how to use this type of interpretation to inform clinical decisions. In contrast, the CEN approach links modalities by directly generating interpretable sample-specific models.

### Sample-Specific Model Parameters

In this work, we identify discriminative subtypes by estimating a basis set of models which can be utilized to generate model parameters specific to each sample. Interest in such *sample-specific* model parameters for cancer analysis has grown in recent years, as increasing evidence has pointed to fine-grained subtypes which do not form discrete clusters [MOP^+^18, BNC^+^15]. Discovering these individualized molecular profiles could lead to substantial advances in the diagnosis and treatment of disease, in addition to refining our biological understanding of the mechanisms behind different diseases. Despite the recent surge of interest in personalized modeling [AYHvdS16, LMP17, MAK^+^07, KTY^+^15, LWJ^+^16, VKVK19], basic statistical challenges remain unanswered and deserve further study. In this work, we contextualize such personalized models with imaging data. We hope that this idea of contextualization can spur further development of such patient-specific modeling.

## B Implementation Details

### Architectural Components

To encode histopathology images, we follow [COS^+^18] and use InceptionV3 [SIVA17] architecture as context encoder. Since this architecture naturally operated on images of size 299×299, we sliced the larger histopathology images into non-overlapping patches of size 299×299, filtering out patches that had more than 25% of background whitespace.

For the interpretable probabilistic model, we used logistic regression (LR) that predicted cancer type from gene expression data. Predictions were computed both on the level of patches (in which case patches from the same slide were assigned the same slide-level labels and gene expression data) and on the level of slides (using majority vote over the corresponding patches). Other choices and hyperparameters are detailed in Section B, and

### Baselines

We compare against both uni-modal and multi-modal baselines. Firstly, we compare against the two uni-modal parts of the CEN architecture: logistic regression on transcriptomic profiles and the convolutional neural network on histopathology images. In addition, we compare performance against two multi-modal baselines. Multi-modal baseline 1 (“Concatenated”) is the same architecture used for the state-of-the-art subtype prediction in [MYA^+^18], which builds a predictor on concatenation of the RNA-seq features and the output of the InceptionV3 network, while multi-modal baseline 2 (“Ensemble”) is an ensemble (i.e., a weighted combination) of the two uni-modal models.

### Data

The data was retrieved from TCGA on April 16, 2019 and preprocessed using the pipeline released by [COS^+^18]. The images are subject to a non-deterministic data augmentation similar to the one implemented in [COS^+^18]. We perform a log-transform and mean-centering of the transcriptomics data and select only those features with a standard deviation greater than 1e-2.

**Table B.1:**
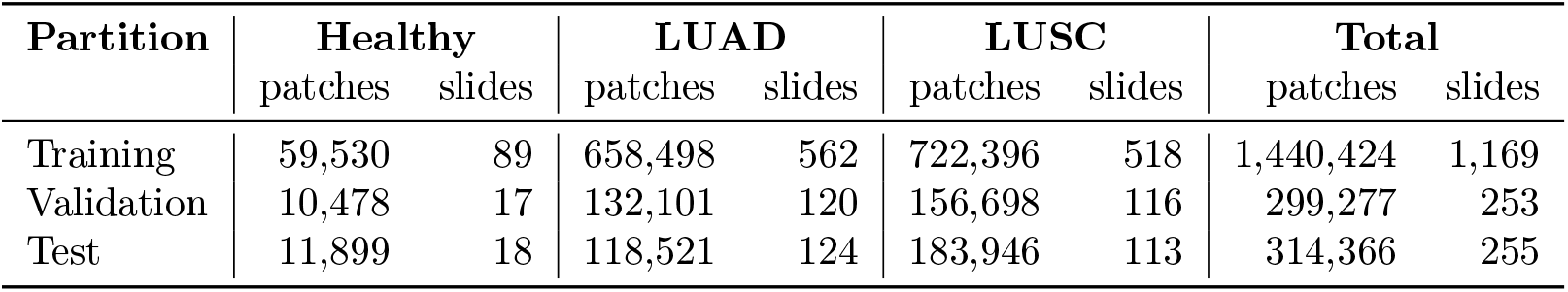
Dataset sizes by counts of slides and patches. The slides are divided into patches.

### Architectures

To the Inceptionv3 archiecture, we added a top dense layer of 128 neurons with dropout (keep probability=0.7). The Contextual Explanation Network parameters have been optimized via a grid-search on the dictionary size along with the regularization of the contextual layer. The final model uses a dictionary size of 32 and *ℓ*_1_ regularization parameter of 1e-5.

### Training

We trained all models for 5 epochs with a batch size of 64 on 1 NVIDIA® Tesla® P100 GPU. The model is optimised using RMSProp with parameters *ρ* = 0.9, momentum = 0.9, *ϵ* = 1.0. The choice of the checkpoints is based on a best validation loss criterion.

## C Extended Results

### C.1 Classification Results

Table C.1 provides the slide-level confusion matrices for all models on the held-out test data. These results, analogous to the accuracy metrics reported in Table 1 of the main text, demonstrate the improved accuracy of CEN over existing models for LUAD/LUSC/Healthy classification.

**Table C.1:** Slide-level confusion matrices for baselines and CEN. Rows correspond to ground truth labels and columns to predictions made by the corresponding models.

|  | CEN |  |  | Logistic Regression |  |  | InceptionV3 |  |  |
| --- | --- | --- | --- | --- | --- | --- | --- | --- | --- |
|  | Healthy | LUAD | LUSC | Healthy | LUAD | LUSC | Healthy | LUAD | LUSC |
| Healthy | 18 | 0 | 0 | 15 | 1 | 2 | 12 | 5 | 1 |
| LUAD | 1 | 117 | 6 | 3 | 115 | 6 | 4 | 102 | 18 |
| LUSC | 0 | 7 | 106 | 0 | 8 | 105 | 1 | 22 | 90 |

|  | Concatenated |  |  | Ensemble |  |  |
| --- | --- | --- | --- | --- | --- | --- |
|  | Healthy | LUAD | LUSC | Healthy | LUAD | LUSC |
| Healthy | 17 | 1 | 0 | 15 | 1 | 2 |
| LUAD | 1 | 115 | 8 | 3 | 115 | 6 |
| LUSC | 0 | 7 | 106 | 0 | 6 | 107 |

### C.2 Intra-tumor Heterogeneity

**Figure C.1:**
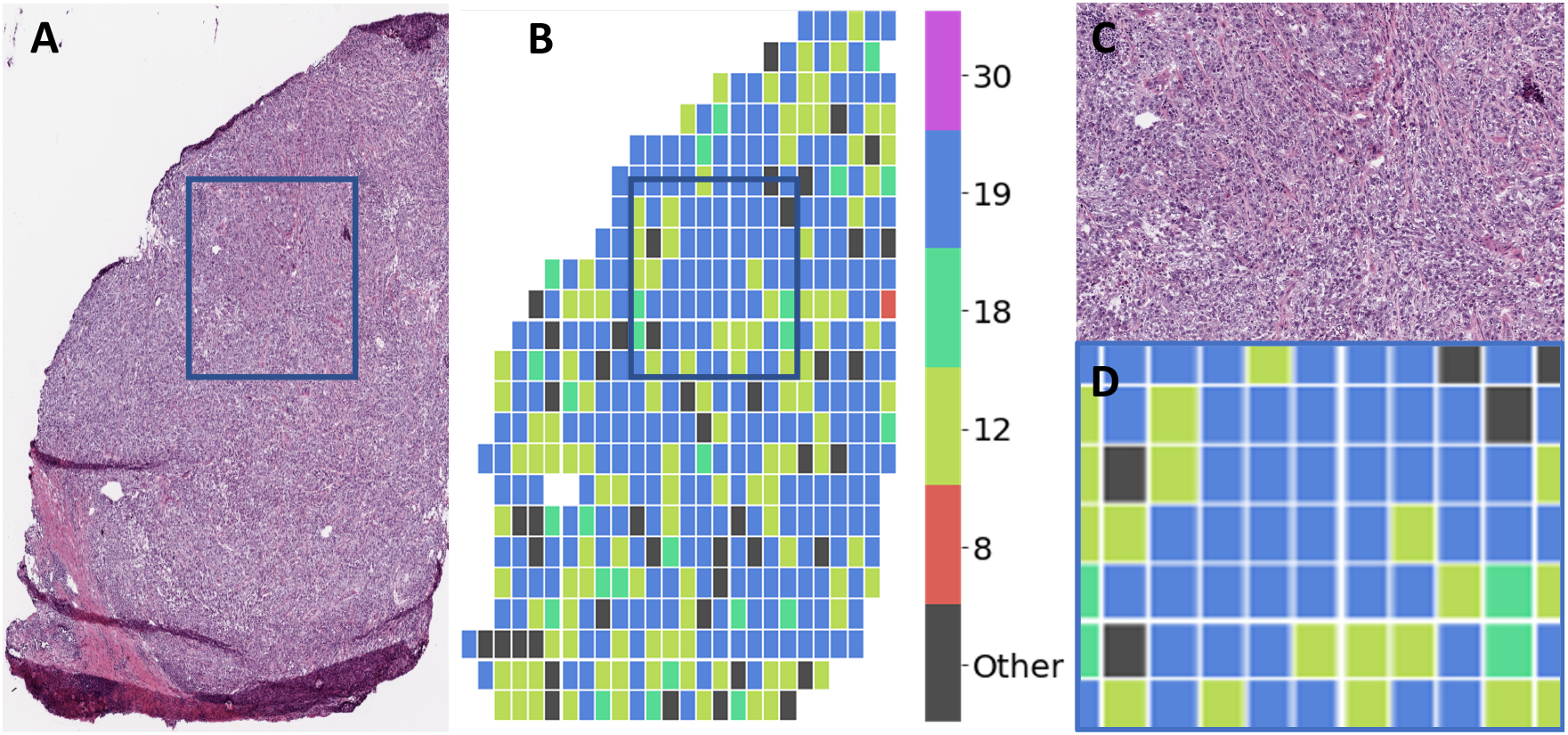
Discriminative subtypes display significant heterogeneity throughout the sample and cluster spatially. Representative results of patch-level subtype prediction for a LUAD sample held out from CEN training. **(A)** shows the tumor whole-slide image. **(B)** displays the archetype which CEN assigned to each patch; for visual clarity, we display only the common archetypes 8, 12, 18, 19, and 30, and group all other archetypes under “other”. **(C)** inset square outlined in blue in panes A and B. **(D)** inset for a matched control sample from healthy tissue surrounding the tumor.

**Figure C.2:**
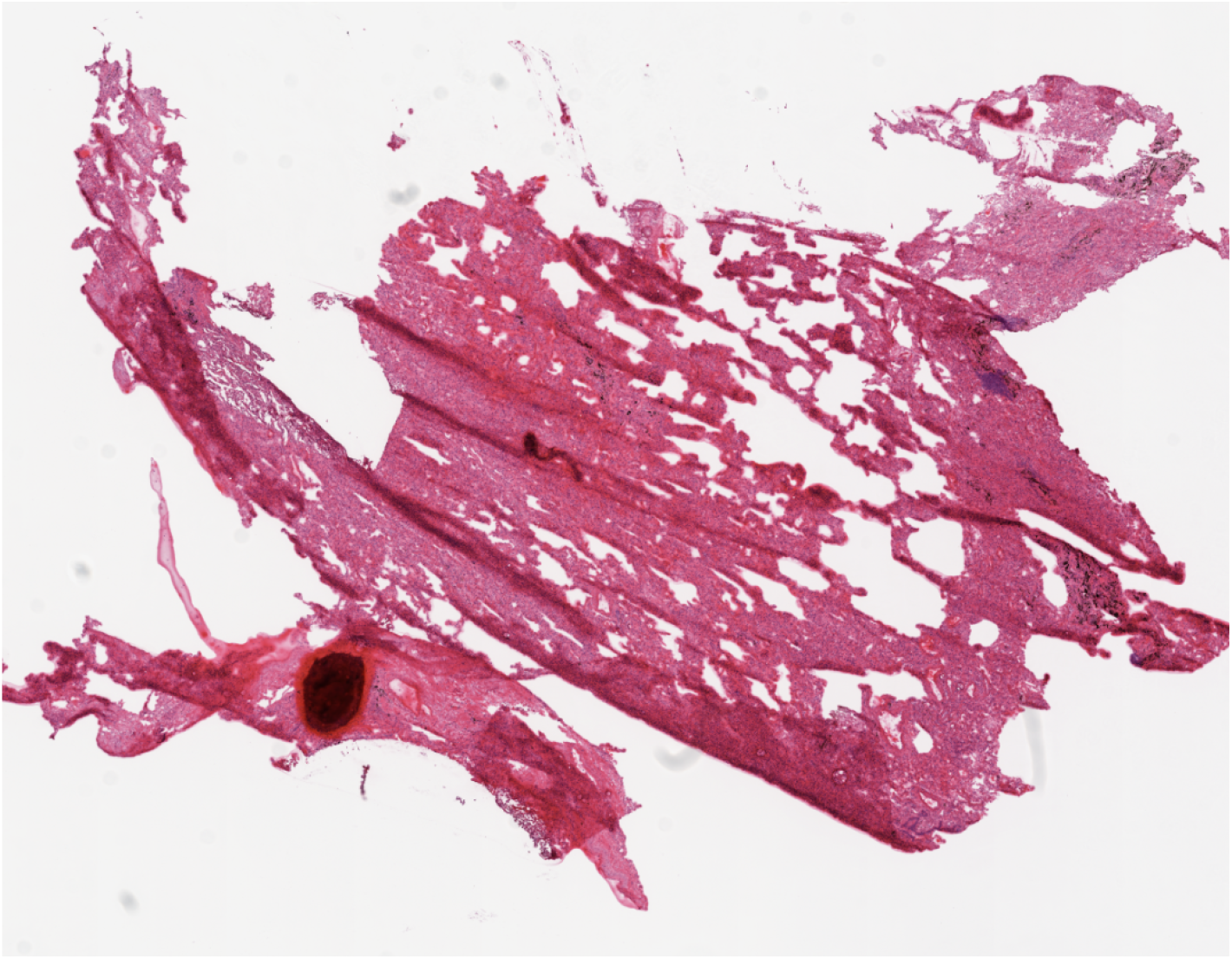
Matching control sample of healthy tissue surrounding the LUSC tumor in Figure 3.

**Figure C.3:**
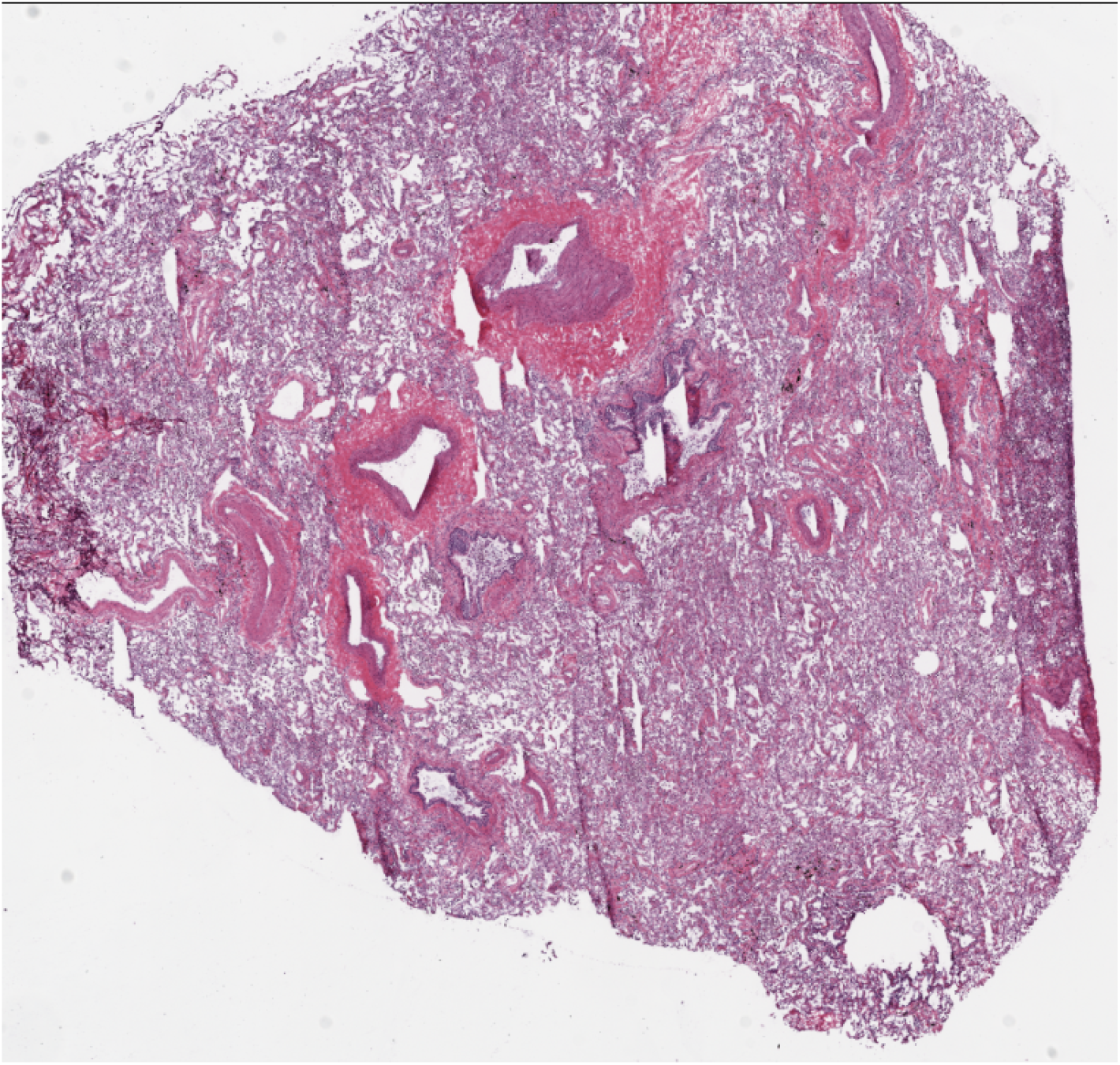
Matching control sample of healthy tissue surrounding the LUAD tumor in Figure C.1.

### C.3 Morphological Patterns of Archetypes

**Figure C.4:**
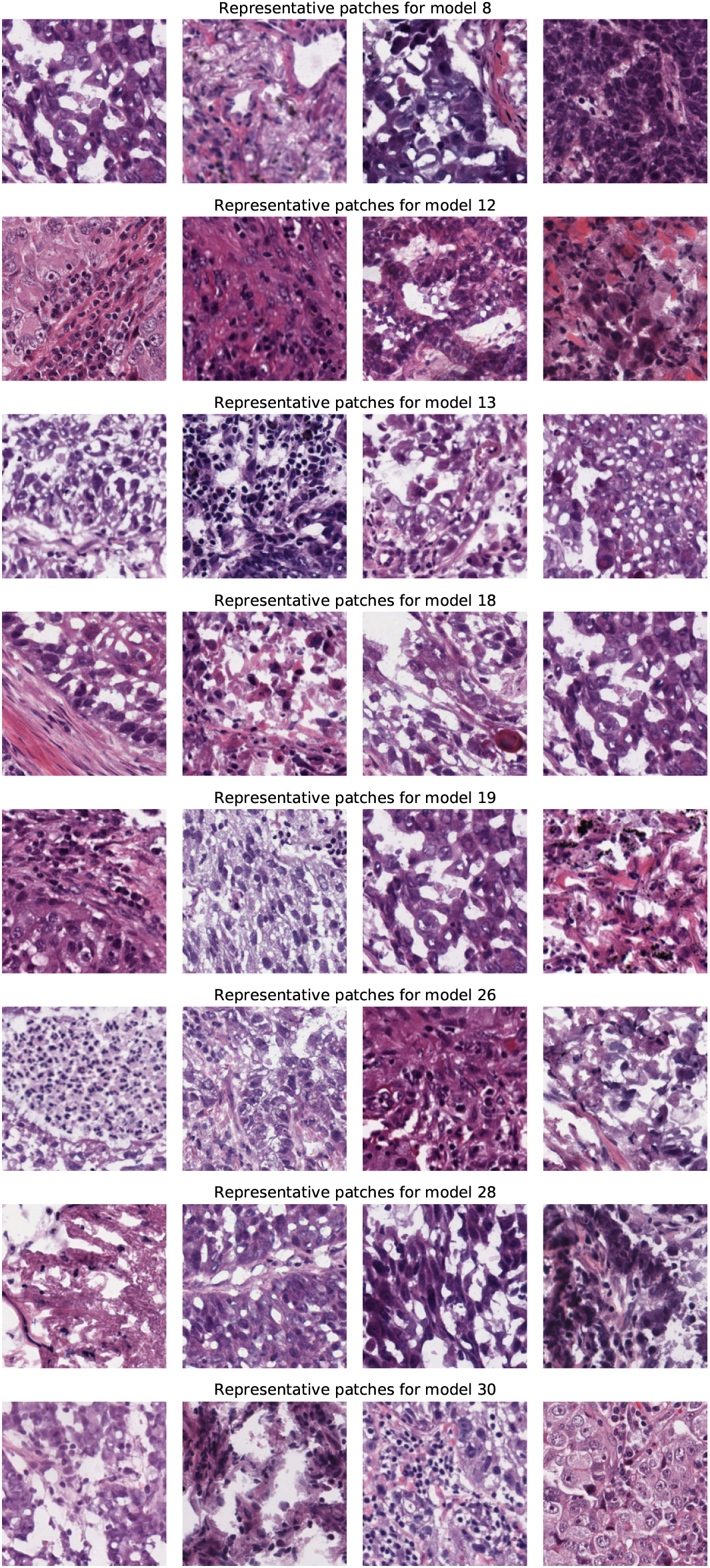
Top four patches that made CEN assign the highest weights to corresponding archetypes.

### C.4 Analysis of the Discriminative Subtyping Hypothesis

Our discussion started with a hypothesis that gene expression data must be interpreted with respect to patient-specific contexts that may correspond to different subtypes. The contextual learning approach we proposed to use in this work allowed us to discover latent subtypes, which we termed *discriminative* since each of the subtypes corresponded to an interpretable linear model that could accurately discriminate between different classes (cancer types in our case) based on transcriptomics.

However, if samples of the same type always mapped to the same linear model, while samples of different types mapped to different models, this would have been troublesome—the biological interpretation of the archetypal coefficients would have been confounded by the assignment of different models to different classes. As shown in Figure C.5, this is not the case: the distribution over the weights assigned to archetypes by the context encoder conditional on the target label are nearly identical across all classes. Thus, indeed linear models that correspond to each of the subtypes are ultimately discriminating between the classes.

While class-conditional distributions over the discovered subtypes are nearly identical, as shown in Figures C.6, C.7, C.8, C.9 different tumor stages do show different distributions of discriminative subtypes. This may reflect the tendency of different subtypes to progress at different rates, or different efficacy of treatment for different subtypes.

**Figure C.5:**
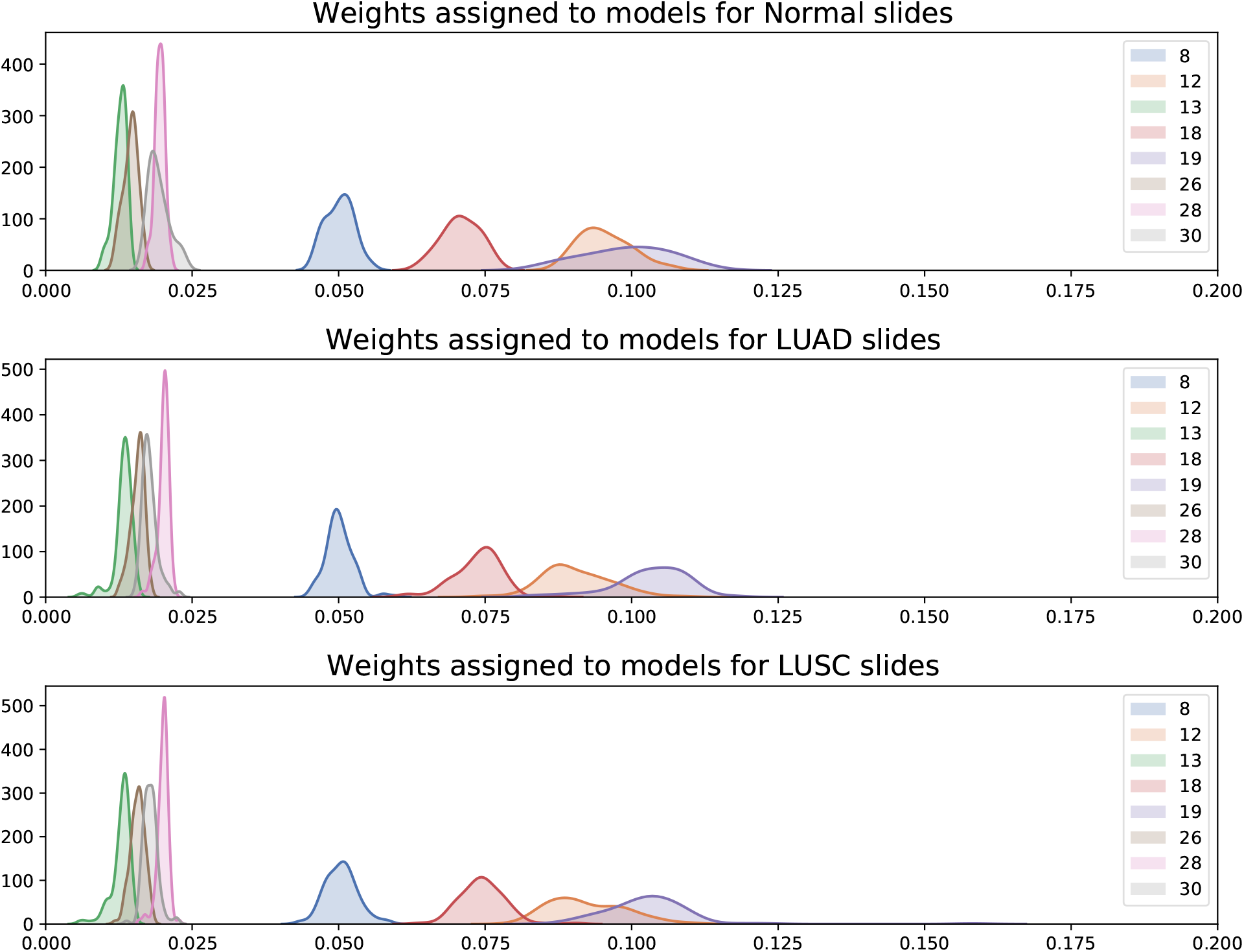
Class-conditional distributions over the archetype weights assigned by CEN, visualized for healthy, LUAD, and LUSC slides. There is only slight variation between distributions of the weights assigned to different archetypes, and the relative ordering between the archetypes is preserved.

**Figure C.6:**
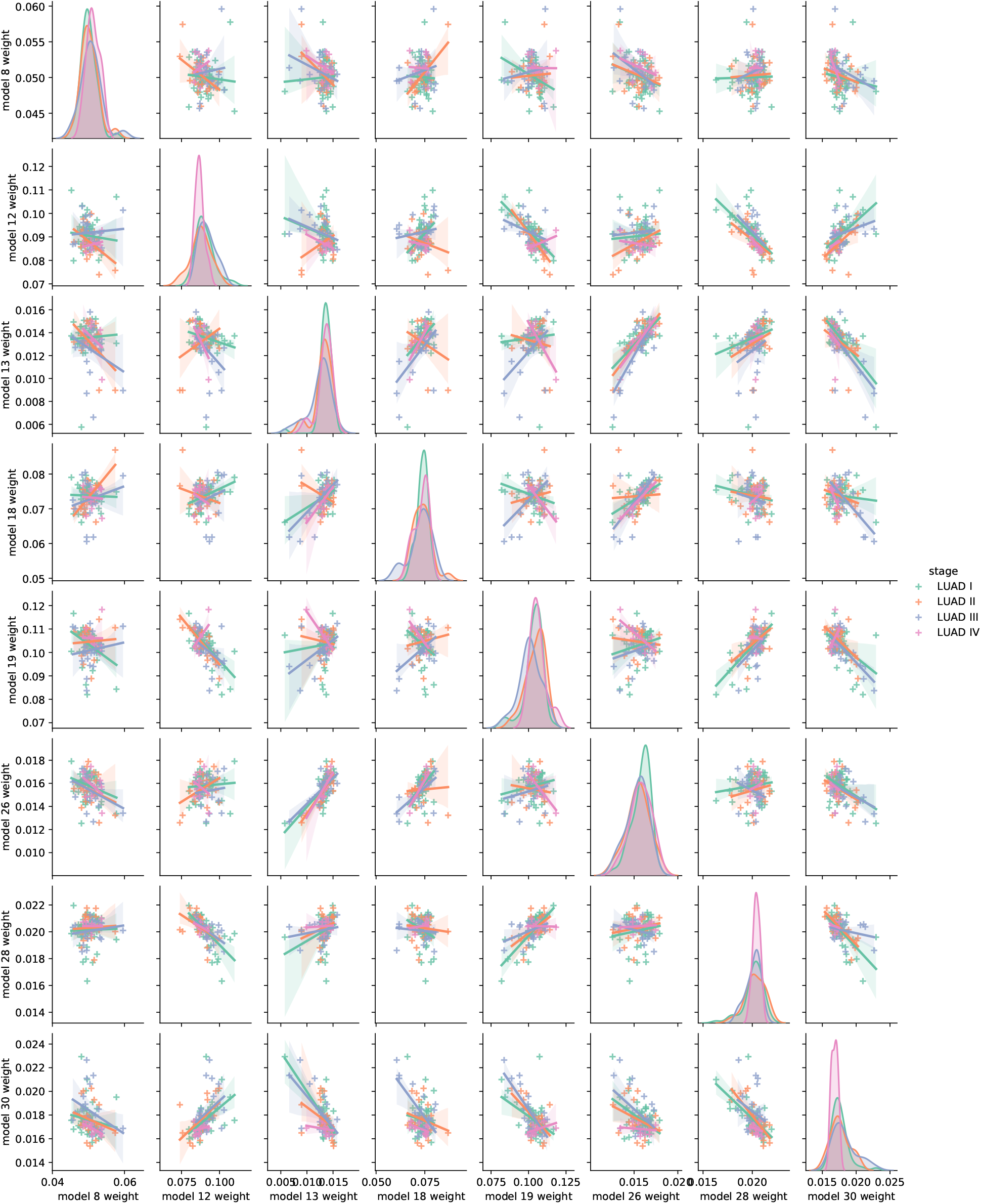
Distribution of weights assigned to 8 different models in the dictionary for different stages of LUAD. Densities on the diagonal indicate variation of the assigned weights for each stage of LUAD. Off-diagonal plots showcase pairwise dependencies between model weights for different stages of LUAD.

**Figure C.7:**
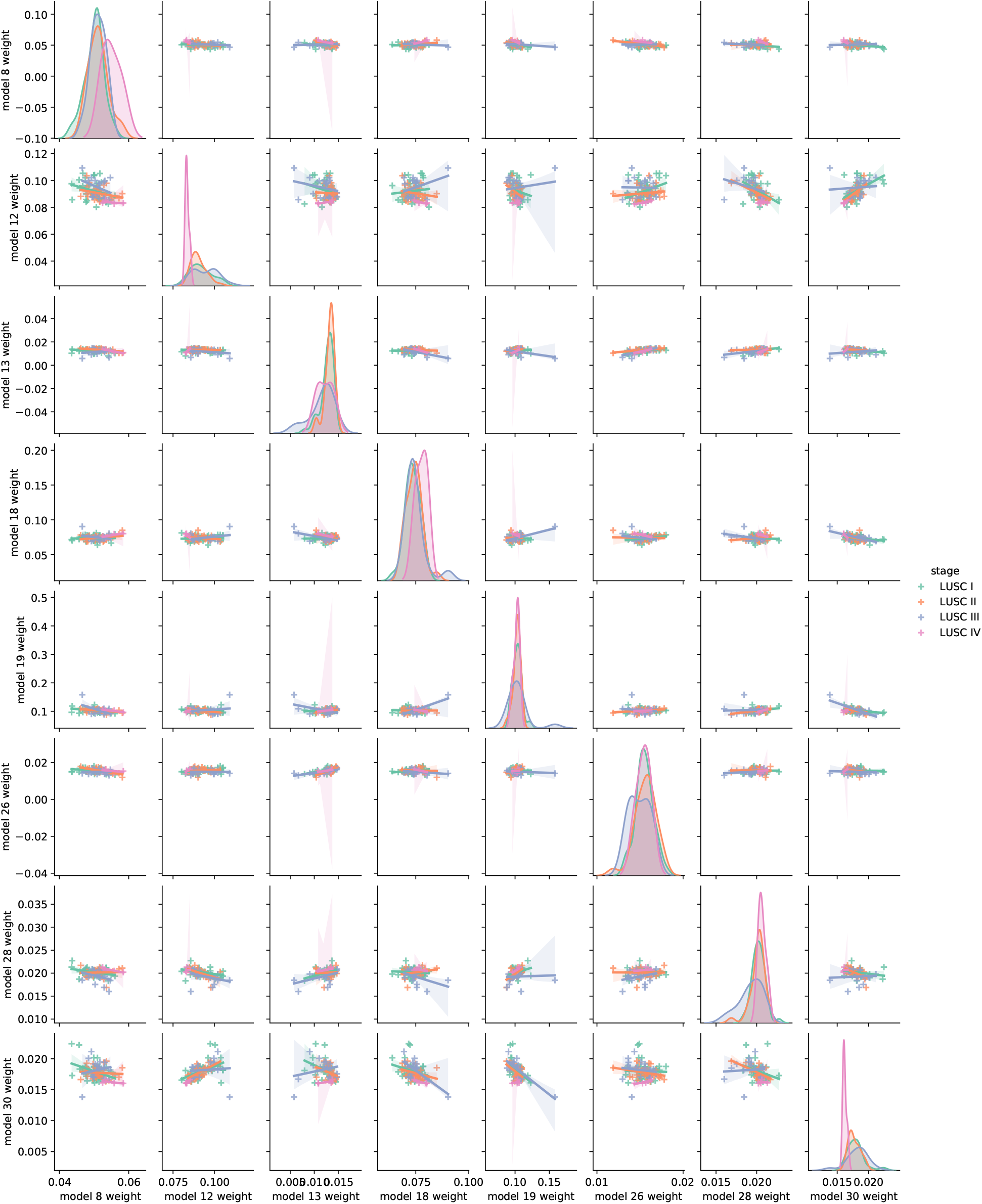
Distribution of weights assigned to 8 different models in the dictionary for different stages of LUSC. Densities on the diagonal indicate variation of the assigned weights for each stage of LUSC. Off-diagonal plots showcase pairwise dependencies between model weights for different stages of LUSC.

**Figure C.8:**
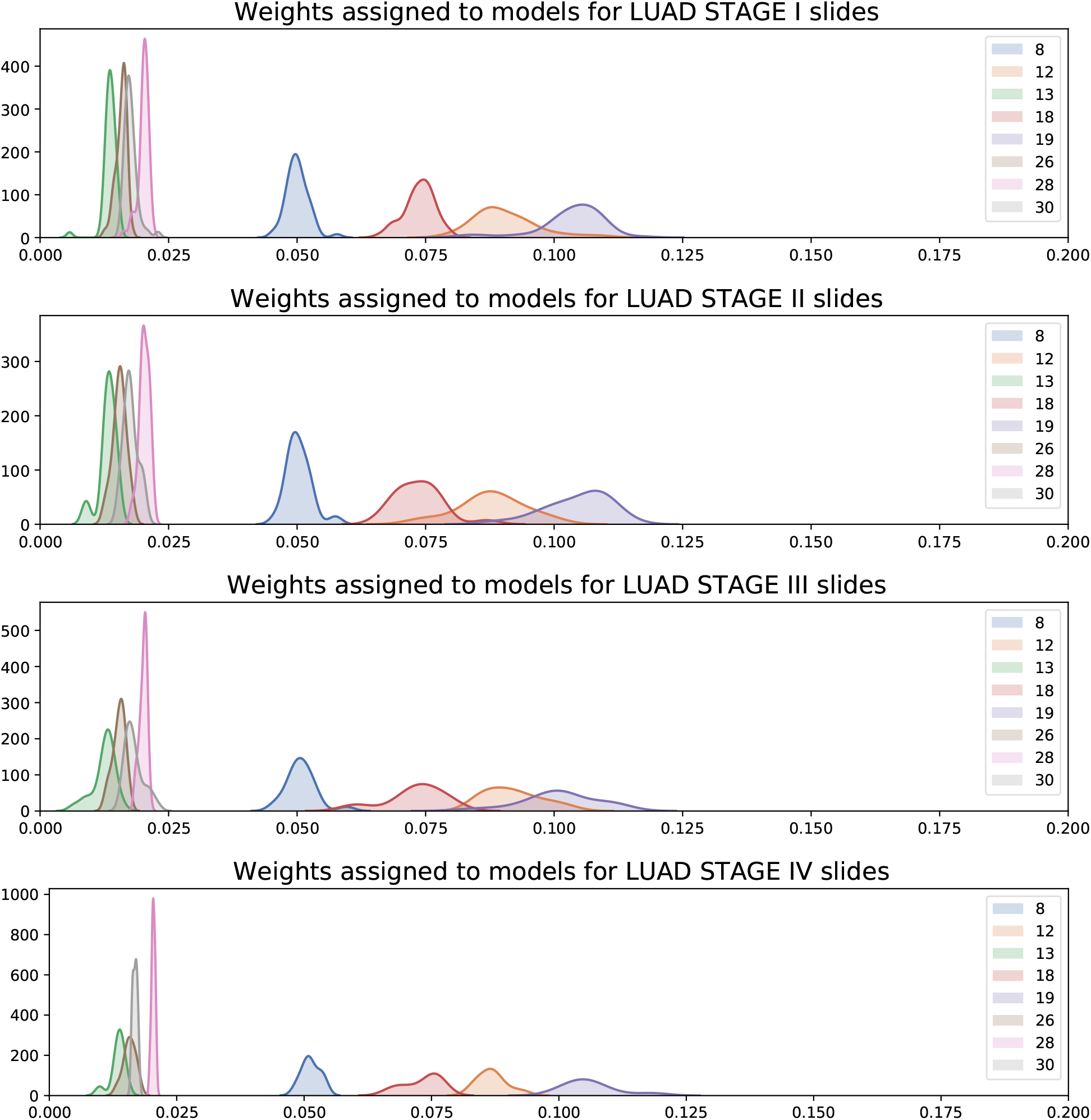
Distributions over the weights assigned to different models in the CEN dictionary visualized for LUAD slides of stages I-IV.

**Figure C.9:**
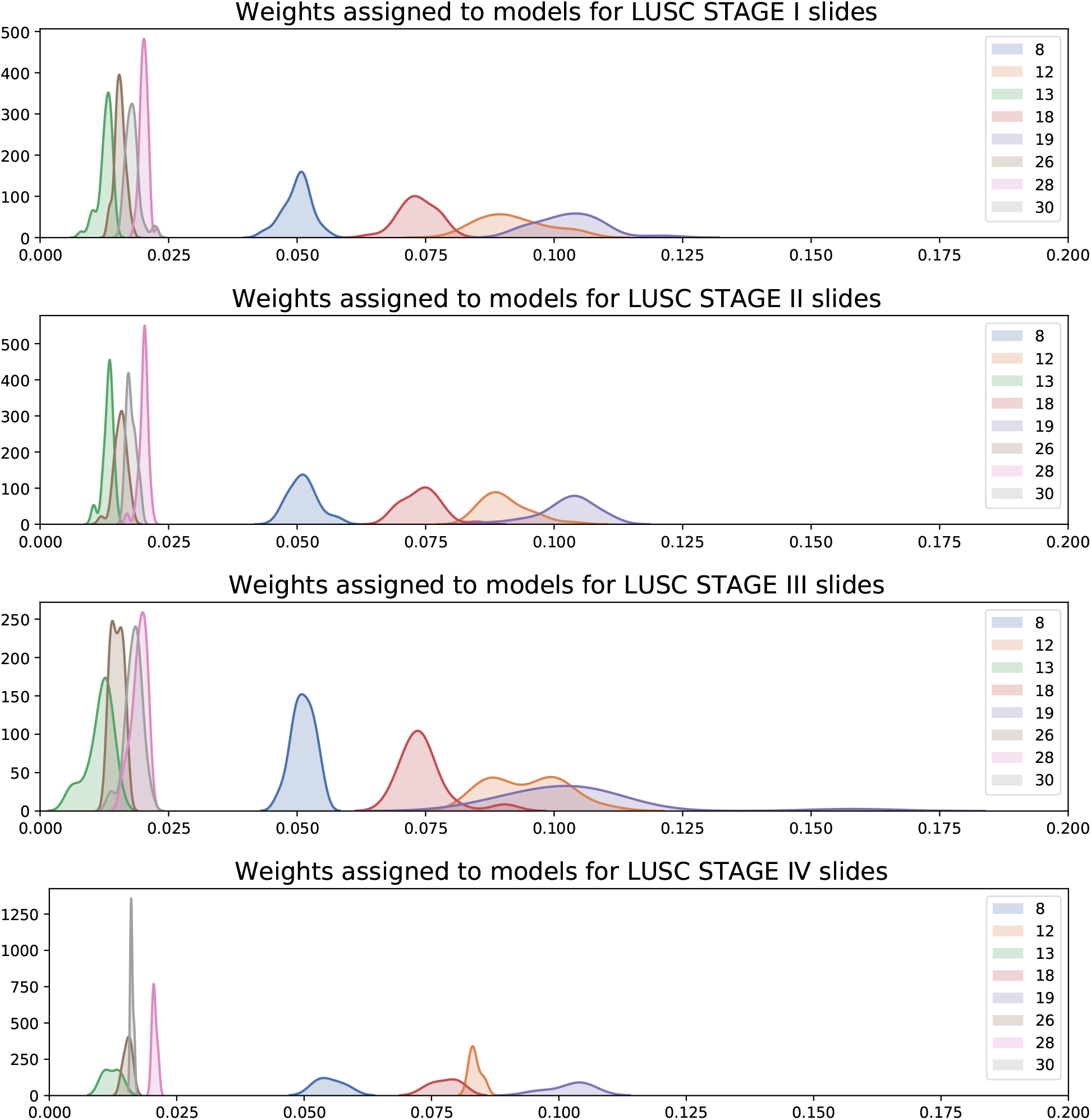
Distributions over the weights assigned to different models in the CEN dictionary visualized for LUSC slides of stages I-IV.

### C.5 Archetype Analysis

#### C.5.1 Enrichment Analyses

**Table C.2:** Terms enriched in LUAD archetypal models (*p <* 0.05).

| Archetype | Term ID | Term Name | P-Value |
| --- | --- | --- | --- |
| 5 | GO:0120038 | plasma membrane bounded cell projection part | 0.038 |
| 7 | TF:M07385_1 | Factor: Lhx2 | 0.008 |
| 11 | GO:0030101 | natural killer cell activation | 0.011 |
| 11 | KEGG:05321 | Inflammatory bowel disease (IBD) | 0.001 |
| 11 | KEGG:04659 | Th17 cell differentiation | 0.003 |
| 16 | CORUM:3075 | UTX-MLL2/3 complex | 0.003 |
| 22 | TF:M00693_1 | Factor: E12 | 0.012 |
| 23 | HPA:008020_13 | cerebral cortex; glial cells[Supported,High] | 0.050 |
| 27 | TF:M01596 | Factor: Gli3 | 0.000 |
| 27 | TF:M07455_0 | Factor: Gli2 | 0.001 |
| 30 | GO:0006259 | DNA metabolic process | 0.017 |

**Table C.3:** Terms enriched in LUSC archetypal models (*p <* 0.05).

| Archetype | Term ID | Term Name | P-Value |
| --- | --- | --- | --- |
| 0 | REAC:R-HSA-6805567 | Keratinization | 0.000 |
| 2 | TF:M02095_0 | Factor: Fra-1 | 0.032 |
| 3 | HPA:050020_02 | spleen; cells in white pulp | 0.022 |
| 6 | GO:0007626 | locomotory behavior | 0.020 |
| 7 | REAC:R-HSA-9006927 | Signaling by Non-Receptor Tyrosine Kinases | 0.035 |
| 8 | TF:M05884_0 | Factor: znf439 | 0.017 |
| 9 | GO:0031435 | mitogen-activated protein<br>kinase kinase kinase binding | 0.011 |
| 12 | GO:0051962 | positive regulation of<br>nervous system development | 0.034 |
| 15 | CORUM:2815 | BRCA1-BARD1-BACH1-DNA<br>damage complex II | 0.029 |
| 16 | REAC:R-HSA-4791275 | Signaling by WNT in cancer | 0.029 |
| 20 | REAC:R-HSA-2173789 | TGF-beta receptor signaling<br>activates SMADs | 0.043 |
| 24 | TF:M08982_0 | Factor: YY1 | 0.020 |
| 26 | GO:0008038 | neuron recognition | 0.010 |

#### C.5.2 Influential Genes in Archetypes

**Table C.4:**
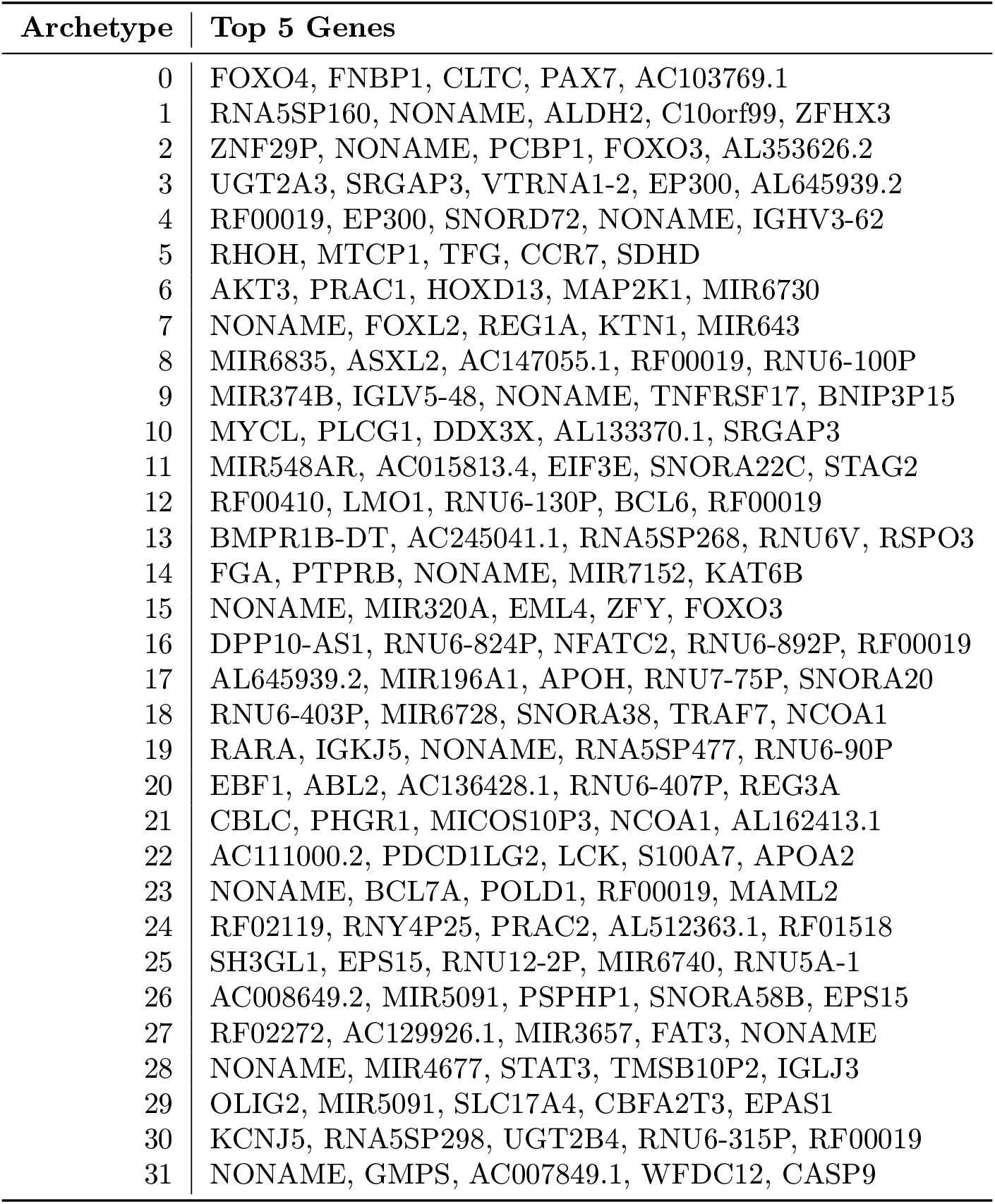
Most influential genes in each case/control archetypal models.

**Table C.5:**
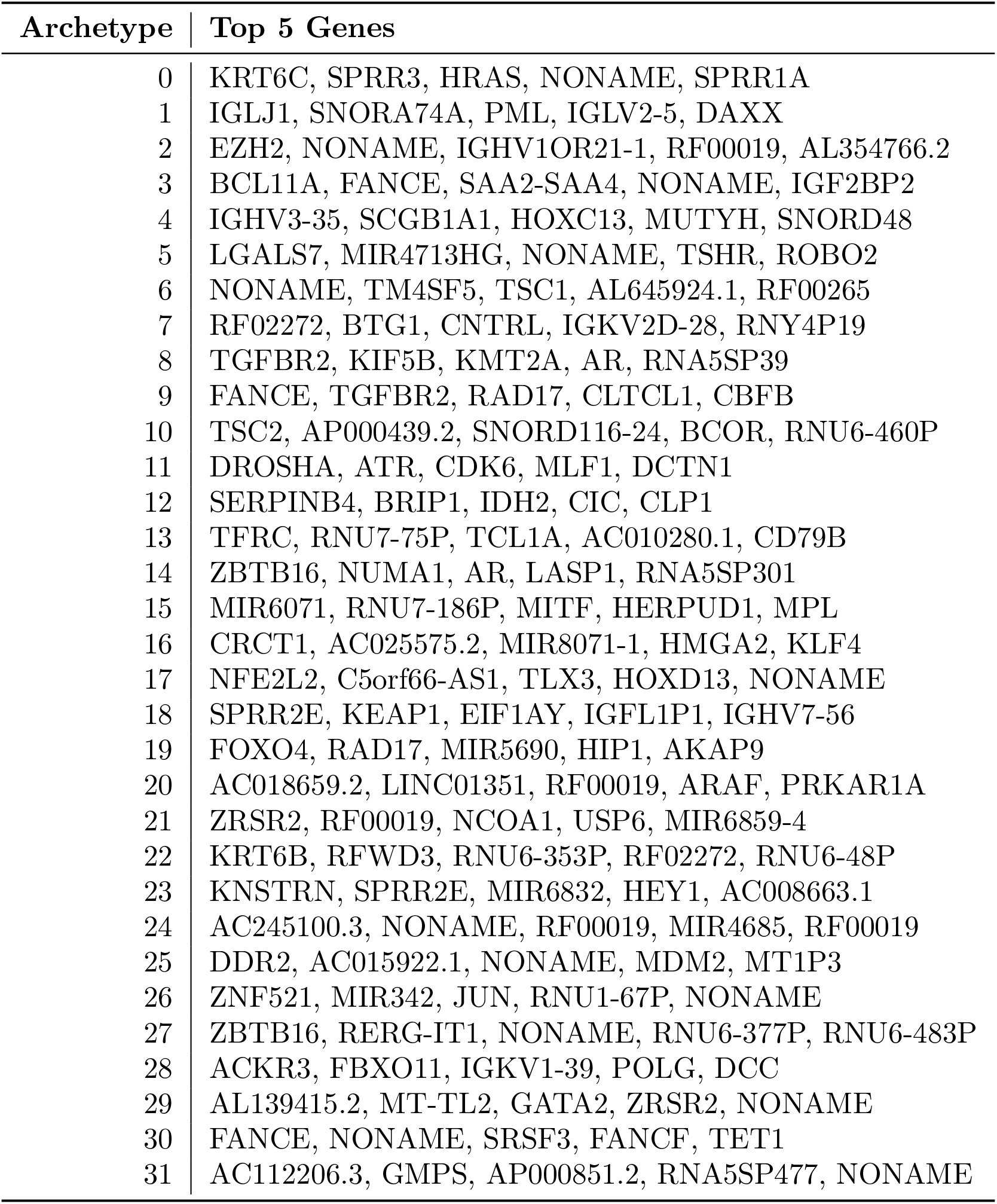
Most influential genes in each LUAD archetypal models.

**Table C.6:**
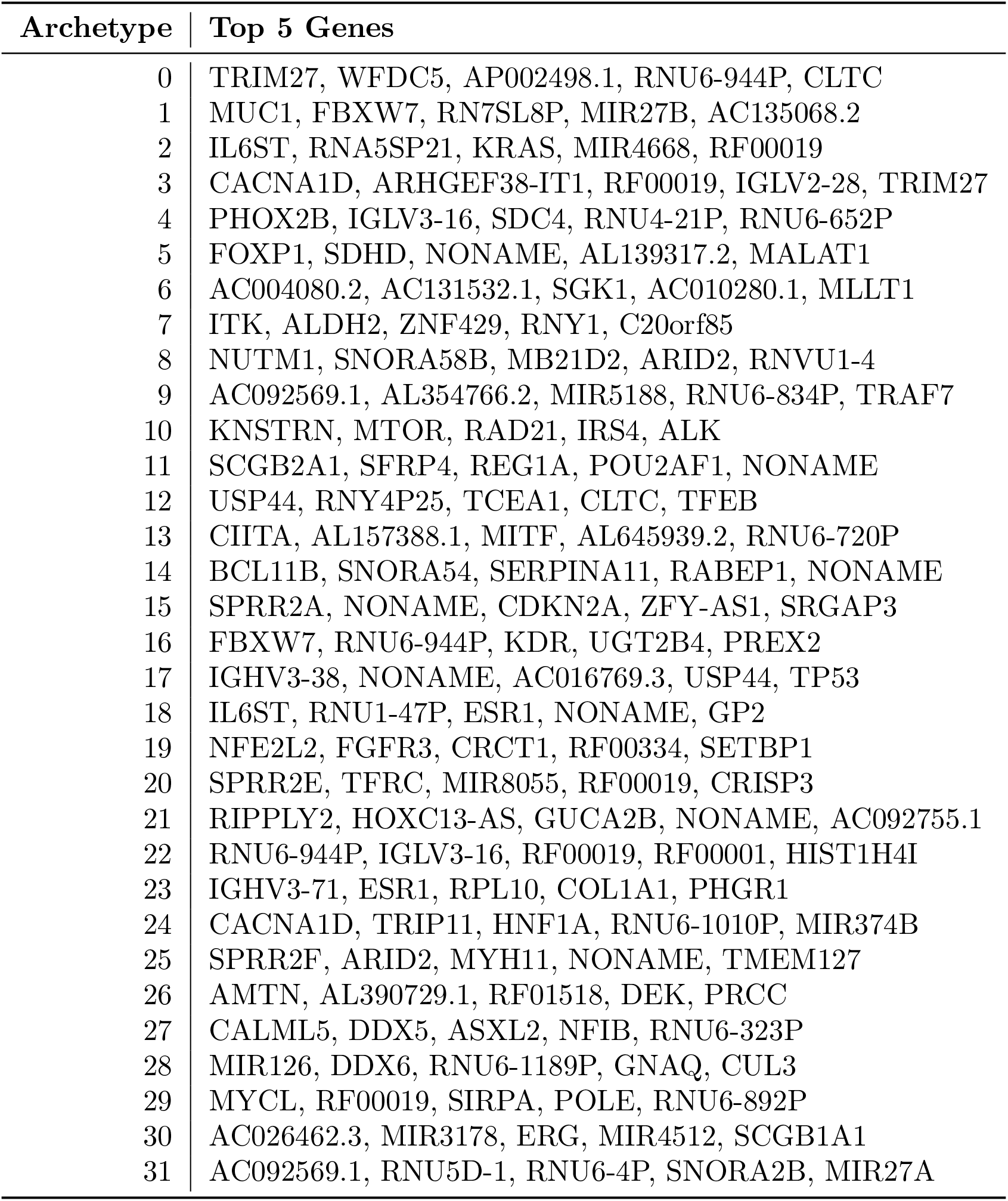
Most influential genes in each LUSC archetypal models.

#### C.5.3 Orthogonality of Archetypes

**Figure C.10:**
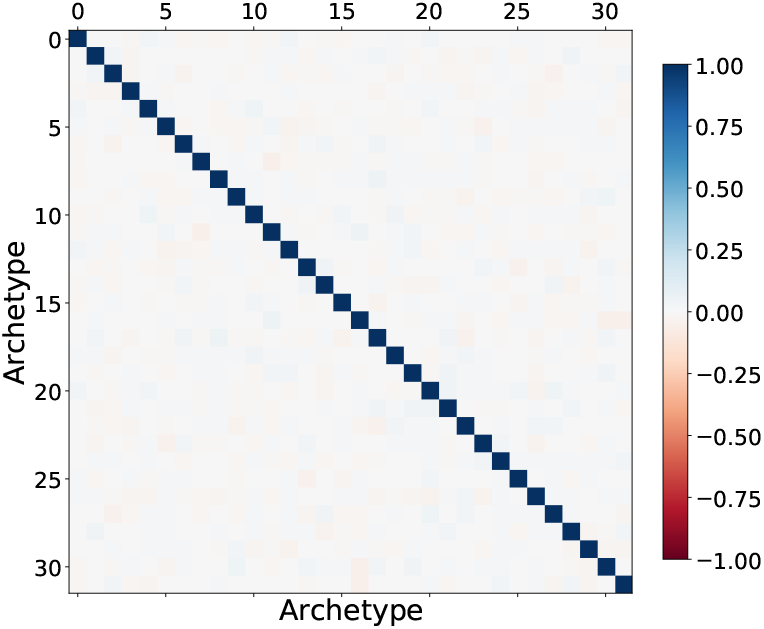
Kendall Tau similarity of ranks of transcripts selected by each entry in the dictionary. All off-diagonal comparisons have similarity less than 0.1, indicating that archetypes are orthogonal.

